# Gender based disparity in performing aortic valve surgery in the united state before availability of percutaneous valve implantation

**DOI:** 10.1101/2023.11.21.23298848

**Authors:** Mohammad Reza Movahed, Arman Soltani, Mehrnoosh Hashemzadeh, Mehrtash Hashemzadeh

## Abstract

**Background:** Aortic valve surgery has been performed increasingly in high-risk patients. The goal of this study was to evaluate this trend based on gender in the United States before the availability of percutaneous aortic valve replacement.

**Method:** The Nationwide Inpatient Sample (NIS) database was utilized to calculate the age-adjusted utilization rate for aortic valve surgery from 1988 to 2011 in the United States using ICD-9 coding for aortic valve surgery

**Results:** A total population of 258, 506 patients underwent aortic valve between 1988-2011 were available for our study over the age of 20. We found that the age-adjusted rate of aortic valve surgery gradually increased from 1988 until 2009 and stabilized thereafter with a persistently higher rate for men. For men age-adjusted rate in 1988 was 13.3 per 100,000 vs. 27.0 in year in the year 2011 per 100,000. For women, the age-adjusted rate in 1988 was 6.07 per 100,000 vs. 11.4 in year 2011 per 100.000).

**Conclusion:** Aortic valve surgery utilization has stabilized in recent years in both genders in the United States. However, this rate has been persistently more than double in men. The cause of this higher utilization in males needs further investigation.

## Introduction

Aortic valve disease, composed of aortic stenosis AS and aortic regurgitation (AR), has become a significant global concern due to the elevated risk of mortality in severe cases when left untreated (1, 2). AS is caused mainly by degenerative calcification of the leaflets, bicuspid aortic valve, or less commonly, rheumatic heart disease, and it also is the most abundant type of valve disorder in the United States, affecting 2-7% of the population over 65 years old(3, 4). AS constitutes more than 40% of cases with native valve disorders, and its prevalence has been roughly equal between men and women(5).

Aortic valve replacement (AVR) has been the mainstay of treatment for severe AS or AR and has demonstrated effectiveness and improved mortality(6). Harken and Braunwald were the first to perform this surgery, and since then, the use of AVR as a standard treatment for severe aortic valve disorders has prevailed(4, 7). The procedure begins through a median sternotomy and extracorporeal circulation, then cardiac arrest is induced using a cardioplegic solution, and AVR is performed(8). The mortality rate for aortic valve replacement in the absence of coronary involvement is around 1-3%(4).

Sex-related differences in aortic stenosis (AS) are significantly overlooked, leading to unequal treatment outcomes and more adverse results for women, such as disproportionately elevated mortality rates(9). As aortic valve surgery has seen a rising trend, this study aims to evaluate this trend in the US based on gender using the Nationwide Inpatient Sample (NIS) database.

## Methods

### Data source

The NIS database, a part of the Healthcare Cost and Utilization Project (HCUP) in the United States, is a publicly available resource widely used by policymakers and researchers. It constitutes about 20% of the sample of all hospitals for adults and features data from around 7 million unweighted and 35 million weighted hospital stays yearly; the NIS is a valuable resource for analyzing healthcare trends, utilization, cost, and outcomes.

#### Sample selection

From 1988 to 2011 in patients over the age of 20 years, NIS database was utilized to calculate age-adjusted utilization rate for aortic valve surgery in the US. ICD-9-CM codes related to aortic valve surgery in the United States, spanning from 1988 to 2011, were accessible for analysis. We specifically employed records where ICD-9-CM was identified as the primary procedure code, denoted explicitly by the code for aortic valve surgery (V43.3). Additionally, we categorized our data into groups based on gender.

## Statistical analysis

Utilizing age-specific weights and rates, we calculated age-adjusted utilization rates for aortic valve surgery from 1988 to 2011. Weighted rates, reflecting the U.S. 2000 standard population, were summed across age groups, and ANOVA trend [M1] analysis was used in our study. The Statistical Package for Social Sciences (SPSS) software was used for this study and data analysis. A p-value of less than 0.05 was accepted as statistically significant.

## Results

Our investigation encompassed a comprehensive cohort comprising 258,506 patients aged 20 and above who underwent aortic valve surgery between 1988 and 2006. The study revealed notable trends in the age-adjusted rates of aortic valve surgery over the specified period. From 1988 to 2009, there was a gradual increase in the age-adjusted rate, which subsequently stabilized until the study ended in 2011. Interestingly, the rate remained consistently higher throughout this duration for male patients. For males, the age-adjusted rate was 13.3 per 100,000 in 1988, and this increased to 27.0 per 100,000 in 2011. In comparison, the age-adjusted rate for females was 6.07 per 100,000 in 1988 and rose to 11.4 per 100,000 in 2011. (Figure 1).

**Figure 1:**
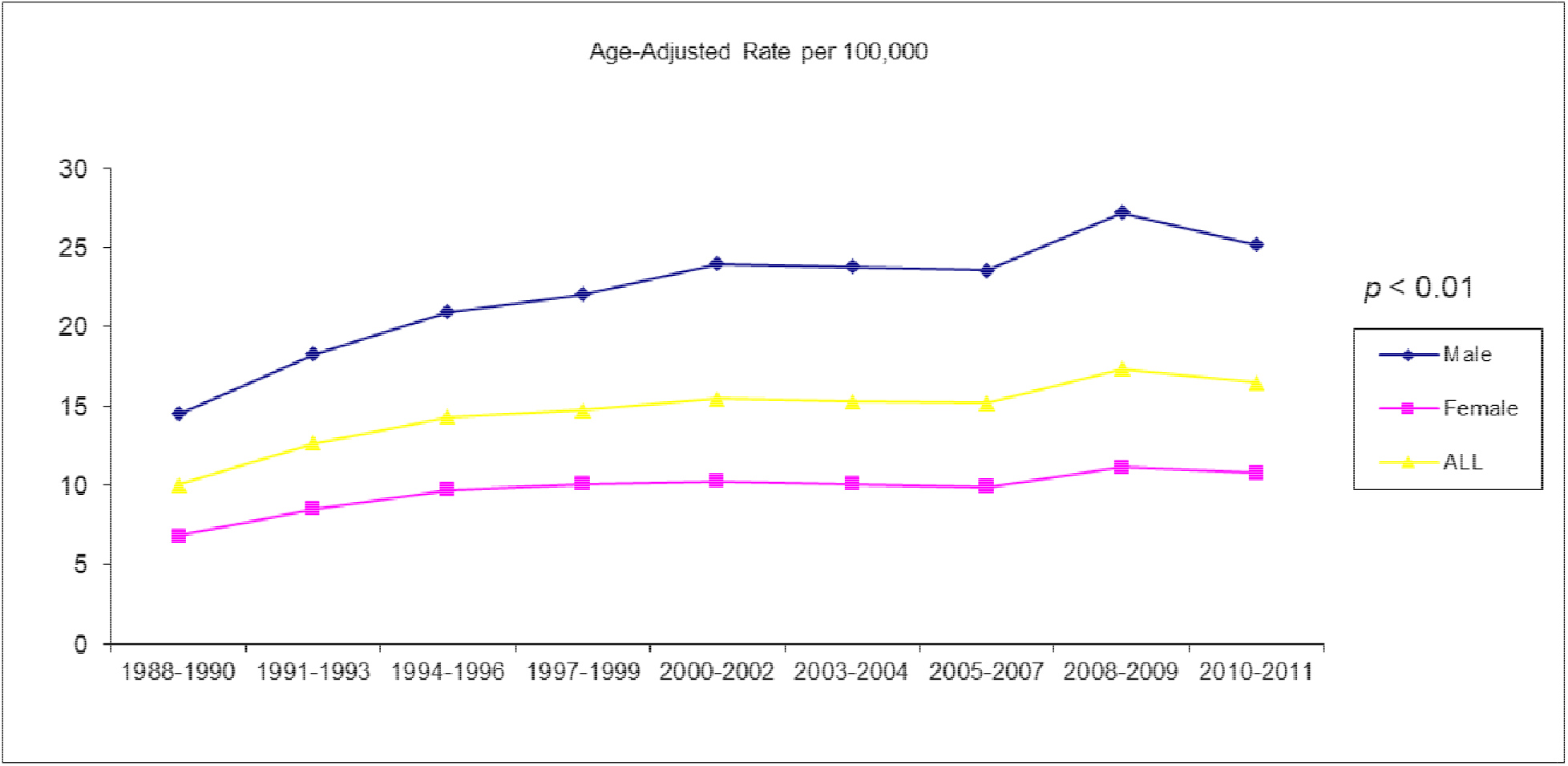
Age-adjusted rate based on gender

## Discussion

We found that the rate of aortic valve surgery performed in the United States has gradually increased in both genders from 1998 until 2000 and stabilized from 2000 until the study ended in 2011 in the area when percutaneous aortic valve implantations were not available. The reason for this inclement and later stabilization of the rate is unknown. Few theories could possibly explain this trend. There are suggestions that improvement in cardiovascular prevention and treatment has led to a lower degree of aortic valve disease in the population(10). Advancements in medicine, a dramatic decrease in post-rheumatic fever, and more effective treatment and management of endocarditis, the underlying causes of stenotic valve development, have paved the way for better control of rheumatic aortic disease(11, 12). Additionally, since aortic valve diseases share similar risk factors with other cardiovascular diseases, such as atherosclerosis (13), improving atherosclerosis with lipid-lowering agents could affect the course of aortic disease and its progression.

Another crucial factor that contributed to stabilizing the rate of valvular surgery performed was the new guidelines for the use of stenotic valve replacement that were published in 1998 by the American Heart Association and American College of Cardiology. According to data published by AHA, for initial diagnosis of aortic stenosis, doppler echocardiography was recommended, as well as regular reevaluation of the patient’s symptoms and progression of aortic stenosis and left ventricle’s function and hemodynamics(13, 14). These evaluations might enable healthcare providers to closely monitor the patient closely, ultimately ameliorating and maintaining the progression of aortic valve diseases.

Prophylaxis was suggested for infective endocarditis, the underlying cause of valvular complications (11, 15). Dramatic improvement in dental hygiene and prophylaxis in 1999 marked a significant advancement in decreasing IE (16). As a result of oral health improvement based on the newly published guidelines about better hygiene and regular brushing and flossing, there was a drastic decrease in oral streptococcal IE. Endocarditis is usually due to the presence of bacteria in intravenous blood, which lodges in the lining of the heart and causes valvular dysfunction. Prevention of aortic stenosis by more effective treatments for IE and the use of prophylaxis antibiotics before and after any dental, GI, and upper respiratory procedures(17) were important factors in stabilizing the rate of aortic valve surgery in 2000 by minimizing the risk of IE. Furthermore, the role of new transcatheter treatments, such as transcatheter aortic valve replacement initially performed in 2000(18), and advances in technology cannot be ignored.

There is increasing awareness of having great outcome statistics around surgeons. This may have led to selection bias to operate only in patients at low risk for bad outcomes. There have been many debates regarding the risks and benefits of aortic valve surgery performed in high-risk patients. Many factors, such as recovery time, mortality rate, risk of infection, and patients’ medical history have played important roles in the trend of aortic valve surgery observed a decade ago(19).

Published data concerning risk factors that contributed to the progression of aortic stenosis and the rate of its progress further led to treatments that managed the stenosis and, therefore, decreased the need for valve replacement and surgery. According to data, smoking, elevated serum calcium, cholesterol level, and male sex were risk factors that increased the rate of aortic stenosis progression(20). We also evaluated this trend based on gender. We found that the rate of aortic valve surgery was much lower in women. Some possible explanations for this pattern could be the dramatic decrease in post-rheumatic fever due to medical advances and fewer valvular complications. Additionally, databases in the US have shown that women’s referral rates to visit a specialist for aortic stenosis or undergoing diagnostic tests have been significantly lower than men’s (21, 22). However, further research is needed to better explain the gender-based di[arity in aortic valve surgery with a lower trend of aortic valve surgery observed in women than men.

### Limitations

The research relied on the utilization of ICD-9 coding, a system that inherently possesses certain limitations.

## Conclusion

Aortic valve surgery utilization has stabilized in recent years in both genders in the United States, probably due to advances in cardiovascular disease prevention. However, this rate has been persistently more than double in men. The cause of this higher utilization in males is not well known.

## Data Availability

Publicly available nIS database

